# Socioeconomic Inequalities and Spatial Clustering in HIV Testing Uptake in Ghana: A Concentration Index and Geospatial Analysis of the 2022 GDHS

**DOI:** 10.64898/2026.08.11.26358504

**Authors:** Abubakar Iddrisu Siddiq, Osman Abdul-Fatawu Iddrisu, Hudu Siddick Abubakar, Sebastina Naa Tawiah Sowah, Solomon Quansah Botchway

## Abstract

**Background:** Though progress has been made towards universal access to diagnosis and treatment, inequalities in HIV testing are a major obstacle to HIV care in the world, as not everyone has enough access to HIV testing services. Progress is being made towards universal HIV testing and treatment, but inequalities in HIV testing are also an important hurdle to HIV care.

**Design:** Cross-sectional secondary analysis

**Setting:** Nationally representative survey across all 16 administrative regions of Ghana.

**Participants:** 22,058 respondents (15,014 women aged 15-49 and 7,044 men aged 15-59) from the 2022 Ghana Demographic and Health Survey.

**Primary outcome measure:** HIV testing uptake, defined as ever having been tested for HIV and received results (binary: yes/no).

**Aim:** The aim of this study was to examine socioeconomic inequalities and spatial clustering in uptake of HIV testing in Ghana in the 2022 Ghana Demographic and Health Survey (GDHS). Specifically, it considered the level of inequality in wealth, the factors related to these inequalities, and the spatial distribution of HIV testing and factors associated with uptake of HIV testing.

**Methods:** The secondary analysis of cross sectional data was done on 22,058 respondents from all 16 administrative regions of Ghana by using descriptive statistics, Erreygers concentration index, Wagstaff decomposition, spatial autocorrelation techniques, and multilevel logistic regression.

**Results:** The findings showed that there was significant pro-rich inequality in HIV testing, with the majority of inequalities observable not being accounted for by the difference in wealth, but in education level. There was a significant geographic clustering with hotspots in the south, and coldspots in the north. HIV testing uptake was significantly predicted by wealth, education, and the age, sex, marital status and being covered by health insurance

**Conclusion:** The study revealed that socioeconomic and geographical inequalities still exist and have significant impacts on uptake of HIV testing in Ghana. It suggests specific programs such as those aimed at disadvantaged communities, geographic areas, greater access to health insurance, and improved community-based HIV testing services to help ensure equitable access.

**Strengths and Limitations:**

1. This study used nationally representative data from the 2022 Ghana DHS, covering all 16 administrative regions, enhancing the generalisability of findings to the Ghanaian population.
2. The combined application of the Erreygers-corrected Concentration Index, Wagstaff decomposition, spatial autocorrelation, and multilevel logistic regression constitute a methodologically rigorous multi-analytical framework rarely applied together in the Ghanaian HIV testing literature.
3. GPS cluster coordinates in DHS data are randomly displaced by up to 5 km in rural areas, which may introduce imprecision in spatial analysis.
4. The cross-sectional design precludes causal inference; directionality of associations between socioeconomic factors and HIV testing cannot be established.
5. Self-reported HIV testing may be subject to recall or social desirability bias, potentially leading to overestimation of testing uptake.

## 1. Introduction

HIV burden remain a pressing global health burden (1). Since the HIV epidemic began, 91. 4 million [73. 4–116. 4 million] people have been infected, and approximately 44. 1 million [37. 6–53. 4 million] have died from HIV-related causes. By the end of 2024, about 40. 8 million [37. 0–45. 6 million] individuals worldwide were living with HIV. The global adult prevalence (ages 15-49) is estimated at 0. 7% [0. 0.6-0. 8%], though this burden varies significantly across countries and regions. The WHO African Region remains the most affected, with nearly 1 in 30 adults (3. 1%) living with HIV, representing over two-thirds of all people living with the virus.

Yet, globally, an estimated 5.46 million people living with HIV remain unaware of their HIV status (2). Achieving the global UNAIDS 95-95-95 targets depends largely on timely HIV testing (3,4), since testing serves as the entry point to diagnosis, treatment, and viral suppression (5,6). Although HIV testing services are provided free of charge in many public health facilities in Ghana, uptake remains uneven across different population groups and regions (7,8). This suggests that the benefits of HIV testing are not reaching all people equally, particularly poorer and underserved populations (9,10)

According to the 2022 Ghana Demographic and Health Survey, 63. 9% of young women (15–24 years) reported ever testing for HIV, and socioeconomic factors such as education, wealth, health insurance, and media exposure significantly predicted screening uptake. Also, spatial clustering shows a clear north-south divide, with southern and coastal regions having significantly higher testing prevalence than northern belts according to the GDHS data (11).

Also, further study from the 2022 GDHS supports the evidence that HIV testing uptake differs by wealth status, education, marital status, employment, residence, pregnancy status, and region according to the 2022 Ghana Demographic and Health Survey (GDHS) (9,12,13). Individuals from richer households are more likely to test for HIV than those from poorer households, while urban residents also tend to have higher testing uptake than rural residents (10,14). Similar inequalities have been observed in antenatal HIV testing, where regional and socioeconomic differences persisted despite improvements in overall testing coverage (15,16). These findings indicate that inequalities in healthcare access continue to influence HIV testing utilisation in Ghana.

Earlier 2014 GDHS data showed only 13% of women and 6% of men had tested in the past 12 months, demonstrating progress but also persistent gaps. For women, the increased likelihood of testing was associated with age (15–39 years), being currently married, attainment of post-secondary education, having only one sexual partner, and residence in regions outside of Greater Accra, like Volta, Eastern, Upper West, and Upper East increased the likelihood of testing. For men, being over 19 years old, attaining post-secondary education, and living in the Upper East region were linked to higher testing rates (17).

While previous studies in Ghana have examined socioeconomic factors associated with HIV testing and others have explored geographic differences separately (10,16,18), few studies have considered both socioeconomic inequality and spatial distribution together. This is important because poverty and place of residence are closely linked (19). Poorer households are often located in underserved communities where access to healthcare facilities, transportation, and health resources may be limited. In addition, geographic clustering of low HIV testing uptake may reflect broader structural inequalities within the health system (20–22).

Understanding not only whether people are testing for HIV, but also who is being left behind and where these disparities are concentrated, is important for improving equitable access to HIV services. This study combined assessment of wealth inequality and spatial clustering using the 2022 GDHS to provide a clearer picture of HIV testing disparities in Ghana. Despite free testing at public facilities, access is driven by wealth, education, and geography and not need.

Early identification of HIV infection for treatment thereof is necessary for the reduction in spread and mortality subsequently; therefore, it is imperative for people to know their HIV status, and this can only be achieved through testing and awareness of HIV status.

Hence, this study examines spatial clustering and its associated drivers of inequality, including education, sex, age, marital status, health insurance, wealth, and media exposure, using the Ghana Health Demographic Survey data for 2022. Graphical representation of the geographical distribution of HIV testing and its associated drivers of inequality is the first step in assisting public health personnel and policymakers to distribute resources allocated to HIV equitably to regions and groups in Ghana with a lower likelihood of testing to help reduce the burden, spread and HIV related mortalities subsequently.

## 2. Design and Methods

### 2.1 Study Design and Data Source

This study was based on a cross-sectional secondary analysis of data obtained from the 2022 Ghana Demographic and Health Survey (GDHS). The GDHS forms part of the global Demographic and Health Survey programme and is implemented periodically to generate nationally representative data on population health, reproductive health, and socioeconomic indicators. The survey was conducted by the Ghana Statistical Service in collaboration with the Ghana Health Service, with technical support provided by ICF International through the DHS Programme.

The 2022 GDHS employed a two-stage stratified cluster sampling design. In the first stage, 618 enumeration areas were selected from the national sampling frame. In the second stage, households were systematically sampled within each selected cluster. The survey covered all sixteen administrative regions of Ghana and was designed to produce reliable estimates at the national, regional, urban, and rural levels.

Data for this study were obtained from the DHS Programme data repository after approval was granted for use of the datasets. Three datasets were utilised. The Individual Recode file (GHIR8CDT) contained information on women aged 15–49 years (n = 15,014), while the Men’s Recode file (GHMR8CDT) included men aged 15–59 years (n = 7,044). Geographic information was obtained from the DHS GPS dataset (GHGE8AFL), which contains cluster-level coordinates. To protect respondent confidentiality, geographic coordinates are randomly displaced by up to 2 km in urban areas and up to 5 km in rural areas. Following the exclusion of respondents with missing information on the outcome variable, the final analytical sample consisted of 22,058 individuals.

### 2.2 Outcome Variable

The outcome of interest was HIV testing uptake. This was derived from respondents’ answers to the question, “Have you ever been tested for HIV and received your results?” Individuals who reported having been tested and having received their results were classified as having utilised HIV testing services and coded as 1. Respondents who had never been tested or had not received their results were coded as 0. The variable corresponded to v781 in the women’s dataset and mv781 in the men’s dataset.

### 2.3 Explanatory Variables

The key explanatory variable was the household’s level of wealth. The DHS wealth index is a principal component analysis of household assets, housing characteristics and access to basic utilities. Those who responded are then classified into five wealth quintiles: poorest, poorer, middle, richer, and richest. Other variables were chosen based on previous empirical research looking at determinants of HIV testing and drawn from the overall social determinants of health framework. These factors included sex (female/male), age group (15–19, 20–24, 25–29, 30–34, 35–39, 40–44, 45–49 and 50–59 for males), education level (no education, primary, secondary and higher), marital status (currently married or living with a partner, formerly married, and never married), residence (urban/rural), health insurance status (none/yes), regular exposure to radio or television (no/yes), and region (city, countryside, and rural areas).

### 2.4 Statistical Analysis

The analysis was carried out in 4 stages in line with the 4 objectives of the study. All analyses were performed considering the complex sampling design of the GDHS. Sampling weights were used to ensure the representativeness of the Ghanaian population and clustering and stratification were used in variance estimation. R software was used for the statistical analysis.

#### Descriptive Analysis

Firstly, descriptive analysis of the study population was performed. To describe HIV testing uptake by categories of the explanatory variables, weighted proportions and corresponding 95% confidence intervals were determined. The survey package in R was used to incorporate survey design specifications and sampling weights were adjusted per the DHS recommendations.

#### Socioeconomic Inequality is measured

The Erreygers-corrected Concentration Index was used to assess socioeconomic inequality in uptake of HIV testing. The conventional concentration index may be difficult to interpret, as it is a binary outcome and it may not be bounded correctly when dealing with HIV testing uptake. The Erreygers correction is a remedy for this and is thus recommended for binary health outcomes. The concentration index was computed for the entire sample and then broken down to sex and place of residence. Positive values indicated that more HIV testing was concentrated among higher income groups, and the negative values indicated that it was concentrated among lower income groups. A Wagstaff decomposition analysis was conducted to provide additional detail on what was causing the inequalities observed. We estimated the effect of each explanatory variable on total inequality by multiplying the elasticity of the variable by its concentration index. With this strategy, the relative importance of various socioeconomic and demographic factors to disparities in HIV testing uptake was identified.

#### Spatial Analysis

HIV testing uptake was explored at the cluster level to gain insight into the geographical distribution of HIV testing uptake across Ghana by calculating the weighted proportion of respondents who reported having ever tested for HIV and received the results within each of the 618 DHS survey clusters. These prevalence estimates were then paired with GPS data in DHS geographic data for the respective locations. To test for any spatial autocorrelation, the Global Moran’s I statistic was used to see if the HIV testing uptake was spatially random or showed significant clustering. A positive value of Moran’s I and a value that is also statistically significant would be interpreted as a non-random spatial pattern, in which HIV testing occurs more often in locations that are close together than in those that are far apart. A non-significant value would be interpreted as a random spatial pattern, in which HIV testing is not more likely to occur near each other than far from each other. Using R and the spdep package, a k-nearest neighbour spatial weights matrix with k = 5 (five neighbours) was created to define neighbourhood relationship among the clusters in the survey. This allowed to ensure that enough neighbouring clusters were connected to each cluster, but on the other hand keep the isolated observations to a minimum. The spatial weights matrix was row standardized to help interpret the local spatial relationships. After the evaluation of spatial dependence at a global level, Local Indicators of Spatial Association (LISA) were calculated based on Local Moran’s I statistics at a local level. This helped identify specific areas with higher than expected HIV testing uptake, as well as areas with lower than expected uptake, relative to the HIV testing uptake of neighbouring areas. This was done as four types of cluster: High–High clusters (hotspots) were those with high prevalence in the cluster with high prevalence in the neighbouring clusters; Low–Low clusters (coldspots) were those with low prevalence in the cluster and in the neighbouring clusters; High–Low clusters were those with high prevalence in the cluster and low prevalence in neighbouring clusters; and Low–High clusters were those with low prevalence in the cluster but high prevalence in the neighbouring clusters. The 5% level was used to determine the statistical significance. The spatial distribution and clustering of HIV testing uptake was plotted using the sf and ggplot2 packages in R.

#### Multilevel Logistic Regression Analysis

The analysis was performed using Multilevel Logistic Regression Analysis. Because of the hierarchical nature of the DHS data, multilevel logistic regression models were used to reflect the clustering of individuals within survey clusters. The first level of analysis was individuals, and the second level was survey clusters. The modelling approach was chosen because people who live within the same cluster might have common social, economic and health system characteristics that may affect the likelihood of HIV testing. An empty (null) model with only a random intercept at the cluster level was first estimated and then the explanatory variables were introduced. The extent of between-cluster variation was quantified using this model and the intraclass correlation coefficient (ICC) was calculated. The ICC was estimated by latent variable method for logistic regression and is defined as the percentage of variance due to differences among clusters. Then, a multivariable model was fit that added wealth quintile and all other explanatory variables as fixed effects and still kept the cluster-level random intercept. This model allowed for simultaneous estimation of individual socioeconomic and demographic risk factors for HIV testing uptake, while also controlling for unobserved contextual risk factors at the cluster level. Maximum likelihood procedures with the use of Laplace approximation were used to estimate the model parameters. Odds ratios (ORs) and their associated 95% confidence intervals (CIs) were presented to measure the association, or strength, of the explanatory variables and HIV testing uptake. Two-sided alpha 0.05 was used to determine statistical significance. The estimates were made nationally representative by using weights for sampling provided by the DHS that were applied throughout the analysis.

### 2.5 Ethical Considerations

This study relied on anonymised secondary data solely from the DHS Programme with a data use agreement. The data sets had no personal identifiers, and no effort was made to identify persons. The Ghana Health Service Ethics Review Committee and the Institutional Review Board (IRB) of ICF International approved the Ethics for the Ghana Demographic and Health Survey (GHS) 2022 before data collection. The analysis of publicly available de-identified data did not require additional ethical approval as this was a component of the present study. All analyses were conducted following the terms and conditions of the data use agreement from the DHS. Geographic positioning was not applied to identify households and individual respondents, but only for spatial analysis at the cluster level.

## 3. Results

### 3.1 Sample Characteristics and HIV Testing Prevalence

A total of 15,014 women (68.1%) and 7,044 men (31.9%) were analysed from the 2022 Ghana Demographic and Health Survey. Forty-seven.6 percent of the respondents reported ever having tested for HIV and received the result. There were socioeconomic disparities in the uptake of HIV testing among the population. The prevalence was gradually increasing by the wealth quintile, from 31.8% (95% CI: 29.6–34.1) among the poorest quintile to 60.8% (95% CI: 58.7–62.9) among the richest quintiles. This is a relative difference of 29 percentage points between the bottom and top of the socioeconomic spectrum, indicating significant differences in access to or use of HIV testing services (Table 1). There were also significant differences found between demographic groups. Females were significantly more likely to have tested for HIV than males, with a prevalence estimate of 57.4% (95% CI: 56.0–58.7) compared to 26.9% (95% CI: 25.2–28.6), respectively. There were also possible geographic differences in HIV uptake with higher uptake among urban residents compared to rural residents (52.0% versus 42.0%). There was a significant positive association between level of education and HIV testing. This proportion also increased gradually with increasing levels of education from 38.1% among those with no education to 72.0% among those with higher education. Likewise, those insured had almost 5 times the odds of being tested for HIV than those who were not (51.4 % v. 26.6 %). There were also differences in age. HIV testing uptake was also significantly higher in adults compared to adolescents, with the largest proportion (two-thirds, 66.9%) of adults (aged 30–34) reporting HIV testing. Among children, however, testing prevalence decreased gently with age after that only 9.3% of adolescents (ages 15-19 years) had been tested for HIV previously. There was large variability across the region within the country. The Eastern Region, Greater Accra Region and Volta Region had the highest HIV testing uptake rates of 55.0%, 54.2% and 53.5% respectively. The lowest prevalence estimates were obtained from the Savannah (25.1%), North East (31.0%) and Northern (31.9%) regions, however. The spatial pattern of these regional differences also indicates important geographies and initial suggests of spatial pattern to be explored in subsequent analyses (Figure 1)

**Figure 1.**
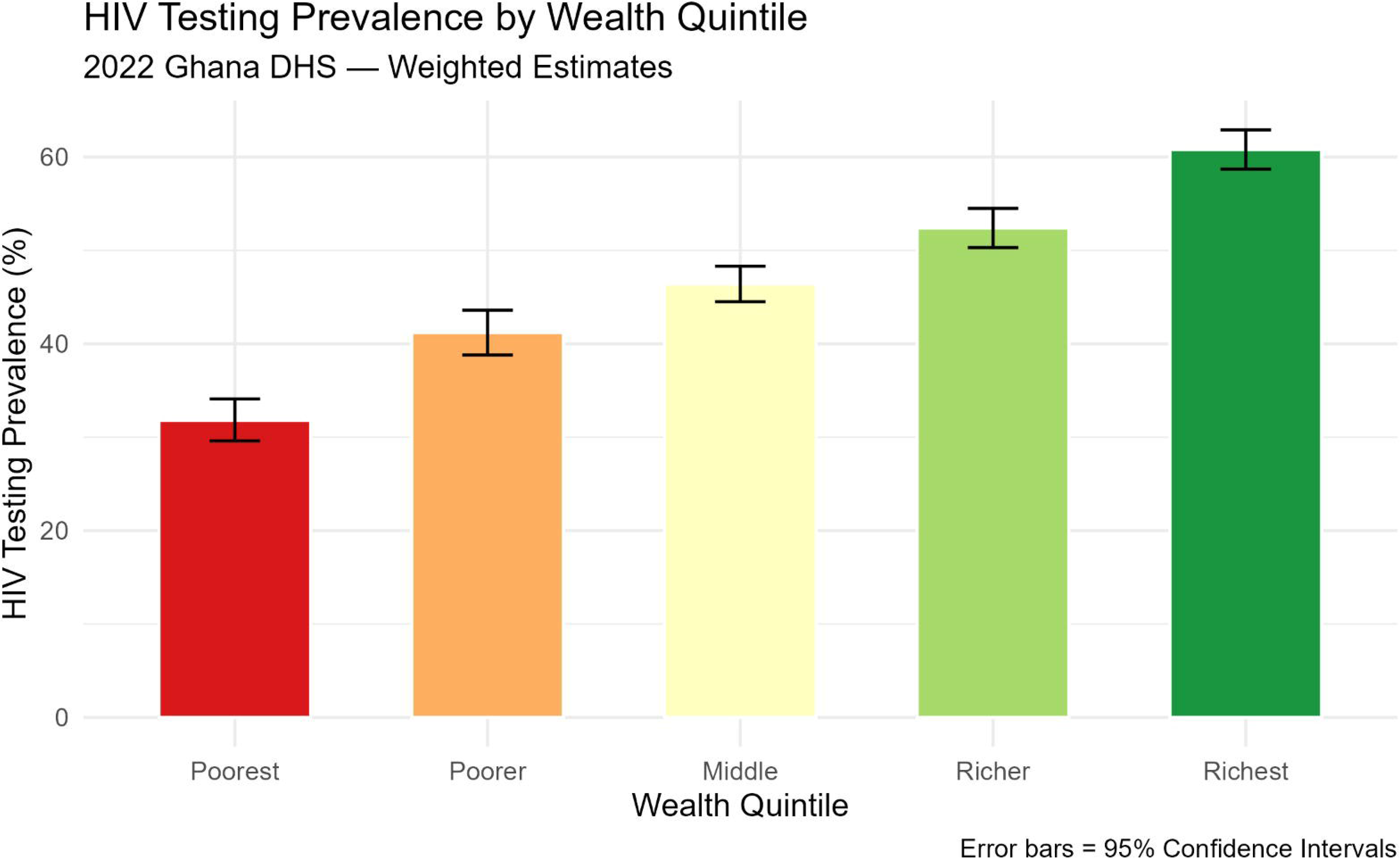
HIV Testing Prevalence by Wealth Quintile, 2022 Ghana DHS. Weighted estimates with 95% confidence intervals. The colour gradient from red (poorest) to green (richest) illustrates the stepwise increase in testing uptake across wealth quintiles.

**Table 1.** Weighted HIV Testing Prevalence by Sociodemographic Characteristics, 2022 Ghana DHS (n□=□22,058)

| Characteristic | Tested (%) | 95% CI Lower | 95% CI Upper |
| --- | --- | --- | --- |
| **Wealth Quintile** |  |  |  |
| Poorest | 31.8 | 29.6 | 34.1 |
| Poorer | 41.2 | 38.8 | 43.6 |
| Middle | 46.4 | 44.5 | 48.3 |
| Richer | 52.4 | 50.3 | 54.5 |
| Richest | 60.8 | 58.7 | 62.9 |
| **Sex** |  |  |  |
| Female | 57.4 | 56.0 | 58.7 |
| Male | 26.9 | 25.2 | 28.6 |
| **Residence** |  |  |  |
| Urban | 52.0 | 50.5 | 53.5 |

| Characteristic | Tested (%) | 95% CI<br>Lower | 95% CI Upper |
| --- | --- | --- | --- |
| Rural | 42.0 | 40.2 | 43.8 |
| **Education** |  |  |  |
| No education | 38.1 | 35.2 | 40.9 |
| Primary | 45.8 | 43.3 | 48.3 |
| Secondary | 45.7 | 44.4 | 47.0 |
| Higher | 72.0 | 69.3 | 74.7 |
| **Health Insurance** |  |  |  |
| No | 26.6 | 24.4 | 28.9 |
| Yes | 51.4 | 50.2 | 52.6 |
| **Age Group** |  |  |  |
| 15–19 | 9.3 | 8.2 | 10.4 |
| 20–24 | 39.5 | 37.2 | 41.9 |
| 25–29 | 61.0 | 58.8 | 63.3 |
| 30–34 | 66.9 | 64.5 | 69.2 |
| 35–39 | 66.5 | 64.1 | 68.8 |
| 40–44 | 60.5 | 57.9 | 63.0 |
| 45–49 | 53.6 | 49.8 | 57.3 |
| 50–59 (men) | 35.5 | 29.4 | 41.6 |
| **Region (selected)** |  |  |  |
| Eastern | 55.0 | 51.7 | 58.4 |
| Greater Accra | 54.2 | 51.4 | 57.0 |
| Volta | 53.5 | 50.1 | 56.9 |
| Ashanti | 51.0 | 47.8 | 54.3 |
| Northern | 31.9 | 26.7 | 37.2 |
| North East | 31.0 | 27.6 | 34.3 |
| Savannah | 25.1 | 19.9 | 30.3 |

*CI = confidence interval. Weighted estimates using DHS sampling weights. — = individual stratum n not separately tabulated.*

### 3.2 Concentration Index and Decomposition

Socioeconomic inequalities in HIV testing uptake were evident in the concentration index analyses. The overall Erreygers-corrected Concentration Index was 0.163 and the overall Standard concentration index was 0.085. The positive value of the index suggests that an unequal distribution of HIV testing services occurred in that higher socioeconomic status populations were more likely to be overrepresented in the distribution of HIV testing uptake, suggesting a pro-rich distribution. There was significant sex-specific socioeconomic inequality, with inequality being higher for men than for women. The Erreyger’s concentration index for men was 0.287 compared to 0.182 for women, indicating that the effect of wealth on HIV testing is stronger among men. In reality, men in better-off households were much more likely to be tested for HIV than men in poorer households. There were also differences noted based on geographic location. The prevalence of HIV testing was higher in urban areas, but there was more socioeconomic inequality in rural areas. The Erreygers concentration index was 0.130 in the rural areas, and 0.096 in the urban areas. This discovery indicates that socioeconomic disadvantage may be a greater obstacle to getting tested for HIV in rural areas where transportation, access to health information and services for HIV testing may be restricted. Overall, the concentration index results suggest that the unequal distribution of HIV testing uptake along the wealth gradient continues to be a significant driver of HIV testing in Ghana. Findings from the persistence of pro-rich inequality across both sex and residence subgroups provide evidence that socioeconomic position remains a factor in the likelihood of using HIV testing services even within population sub-groups that vary in terms of their demographic and geographic characteristics (Figure 2 and Table 2).

**Figure 2.**
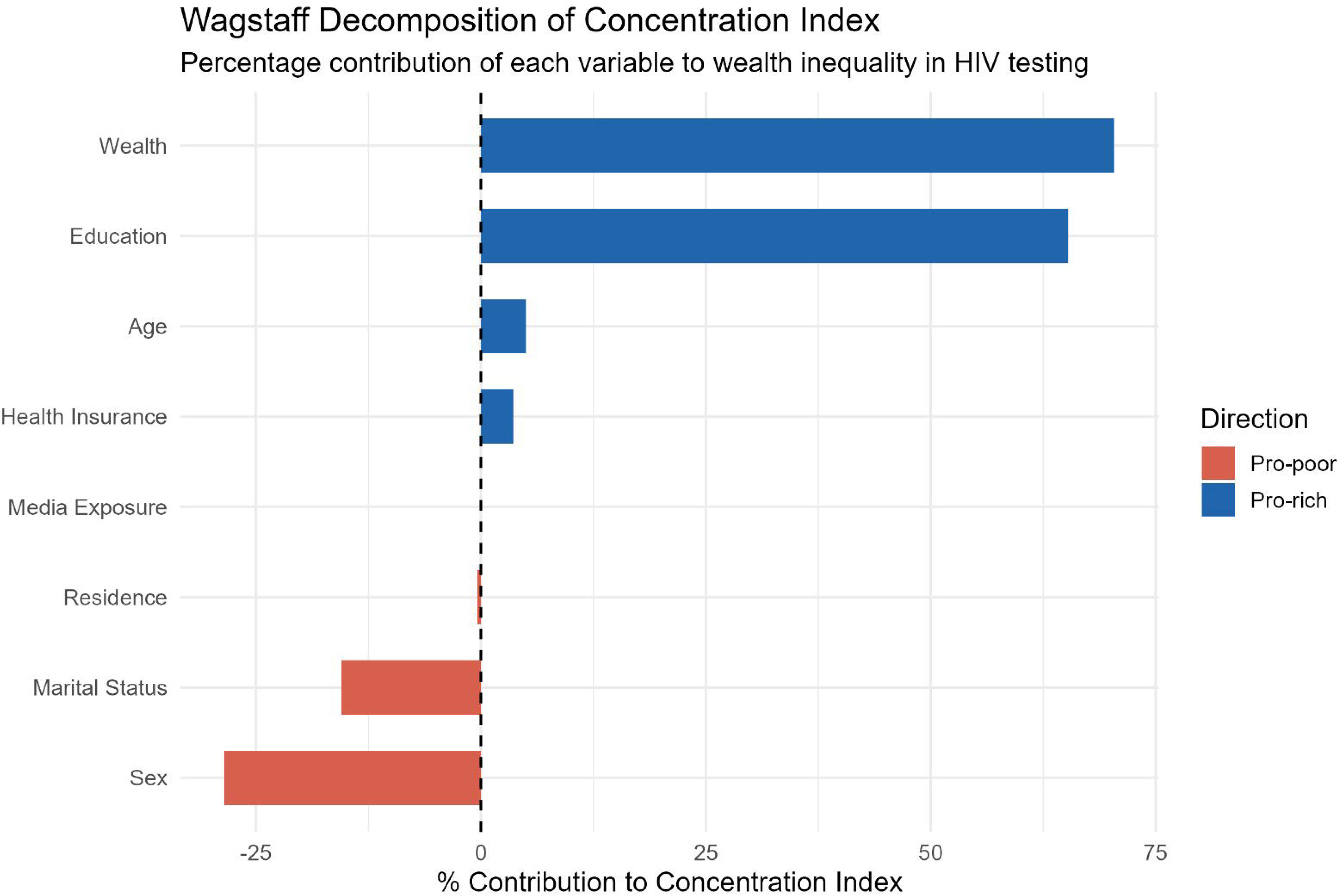
Wagstaff Decomposition of the Concentration Index of HIV Testing Uptake, 2022 Ghana DHS. Bars extending to the right (blue) indicate pro-rich contributions; bars extending to the left (red) indicate pro-poor contributions. Wealth and education are the dominant pro-rich contributors; sex and marital status partially offset the inequality.

**Table 2.** Erreygers-Corrected Concentration Index by Sex and Residence, 2022 Ghana DHS.

| Subgroup | Mean Prevalence | Standard CI | Erreygers CI |
| --- | --- | --- | --- |
| **Overall** | 0.476 | 0.085 | **0.163** |
| Women | 0.574 |  | 0.182 |
| Men | 0.269 |  | 0.287 |
| Urban | 0.520 |  | 0.096 |
| Rural | 0.420 |  | 0.130 |

*All CI values > 0 indicate pro-rich inequality. Erreygers correction applied for binary outcome variable.*

In-depth analysis of the decomposition results showed that the key determinants of inequality in uptake of HIV testing were socioeconomic position and education. The largest contributor was the wealth quintile, which explained 70.4% of the inequality observed. The results indicate that the inequality in household economic resources is still a significant factor in access to and uptake of HTS services in Ghana. Education was the next most important factor, accounting for an extra 65.3% of the overall level of inequality, which shows that education has both an effect on awareness and understanding regarding HIV-related health information and a close linkage to socioeconomic advantage. The results of this regression showed that together, income and education accounted for a significant amount of the inequality in HIV testing uptake, indicating that these two factors are related to each other. Pro-rich inequality was mediated by comparatively small but significant effects of age and health insurance. The proportion reporting HIV testing was higher among older individuals than among younger ones and accounted for 5.0% of the overall inequality. Likewise, health insurance coverage explained the participation of 3.6%, which is consistent with a greater relationship with medical care services, including HIV testing among insured people. Insurance status was more prevalent in higher socioeconomic groups and thus further narrowed HIV testing to those with higher socioeconomic status. Sex and marital status, on the other hand, had negative influences on the concentration index and thus lowered the level of pro-rich inequality. Sex was the biggest equalising factor identified in the analysis with a contribution of −28.5%. This is due to the significantly higher testing prevalence rate among women, which is mainly attributable to routine HIV screening in antenatal and other reproductive health services. The higher proportion of women in the test group, across all classifications of wealth, helped to reduce some of the wealth related inequalities. The effect of marital status was also negative, with inequality decreasing by −15.5%, indicating that inequality in testing rates between marital groups helped to smooth out the inequalities in overall testing rates. In contrast, place of residence and media exposure had relatively little influence on the concentration index. The contribution of these factors is small, suggesting that while they might have an impact on individual HIV testing behaviours they are relatively modest when it comes to explaining the inequalities in the population attributable to wealth. The decomposition analysis shows that most of the socioeconomic inequalities in HIV testing uptake lay in the structural differences of wealth and education and not with geographic residence or access to media (Table 3).

**Table 3.** Wagstaff Decomposition of the Concentration Index of HIV Testing Uptake, 2022 Ghana DHS.

| Variable | $\beta$ | Mean | Elasticity | CI<br>(variable) | % Contribution |
| --- | --- | --- | --- | --- | --- |
| Wealth | 0.038 | 3.163 | 0.249 | 0.249 | **70.4** |
| Education | 0.114 | 2.696 | 0.642 | 0.090 | **65.3** |
| Age | 0.040 | 3.630 | 0.304 | 0.015 | **5.0** |
| Health Insurance | 0.130 | 1.850 | 0.504 | 0.006 | 3.6 |
| Residence | 0.001 | 1.438 | 0.003 | -0.111 | -0.4 |
| Media Exposure | 0.004 | 1.343 | 0.010 | 0.003 | 0.0 |
| Marital Status | -0.148 | 1.866 | -0.575 | 0.024 | -15.5 |
| Sex | -0.282 | 1.309 | -0.769 | 0.033 | -28.5 |

*$\beta$ = regression coefficient from weighted linear probability model. Positive % contributions indicate pro-rich inequality; negative indicate pro-poor. Variables ordered by absolute percentage contribution.*

### 3.3 Spatial Distribution and Clustering of HIV Testing

HIV testing uptake was found to differ significantly by geographic area, within the 618 DHS survey clusters in Ghana. The proportion tested for HIV varied widely across clusters from very low to clusters where a significant portion of residents reported being tested for HIV (Figure 3). The spatial distribution of these differences indicated that they were not randomly distributed across the country but rather appeared to have a spatial pattern. Further evidence was provided by the Global Moran’s I statistic, which gave a value of 0.387 (p < 0.001). A positive and statistically significant Moran’s I suggests that there is high spatial autocorrelation, which implies that neighbouring clusters were inclined to have the same HIV testing uptake. In a nutshell, clusters with high testing prevalence were more likely to be found near other high testing prevalence clusters, and clusters with low testing prevalence were more likely to be found near other low testing prevalence clusters. These results do not support the null hypothesis of spatial randomness and indicate that spatial and contextual factors might affect HIV testing patterns in the country. To gain additional insight into the areas of significant clustering, local Indicators of Spatial Association (LISA) analysis was used. Forty-two clusters were determined to be statistically significant High–High clusters, where HIV testing uptake is a geographic hotspot. The hotspots occurred mostly in the south, mainly in the Greater Accra, Eastern and Volta regions. In these clusters, there were also high levels of HIV testing, surrounded by neighbouring clusters with similarly high levels of HIV testing, suggesting the presence of favourable environments for HIV testing utilisation. On the other hand, 65 clusters were identified as Low–Low clusters, which are coldspots with a low and sustained uptake of HIV testing. Low prevalence clusters were seen to be mainly in the Savannah, Northern and North East regions and surrounded by neighbouring regions with similarly low prevalence testing. The higher prevalence of coldspots in the northern part of Ghana seems to indicate geographical disadvantages such as limited access to healthcare, lower socioeconomic development, and smaller availability of HIV-related services. A few spatial outliers were identified, as well as hotspots and coldspots. Six clusters were termed as High–Low outliers that consist of relatively high testing uptake areas, with low testing uptake neighbours and ten clusters were termed Low–High outliers that consist of pockets of low uptake areas, within areas of high testing uptake. Most clusters (80.1%) had no significant clustering. Put together, these results suggest a definite spatial gradient of the uptake of HIV testing in Ghana. Hotspots in the southern regions and coldspots in the northern regions indicate a stark north–south access to and/or utilisation gap in HIV testing services. The geographic distribution corresponds to other socioeconomic and health system inequities reported nationally, and underscores the need for place-based interventions to increase HIV testing.

**Figure 3.**
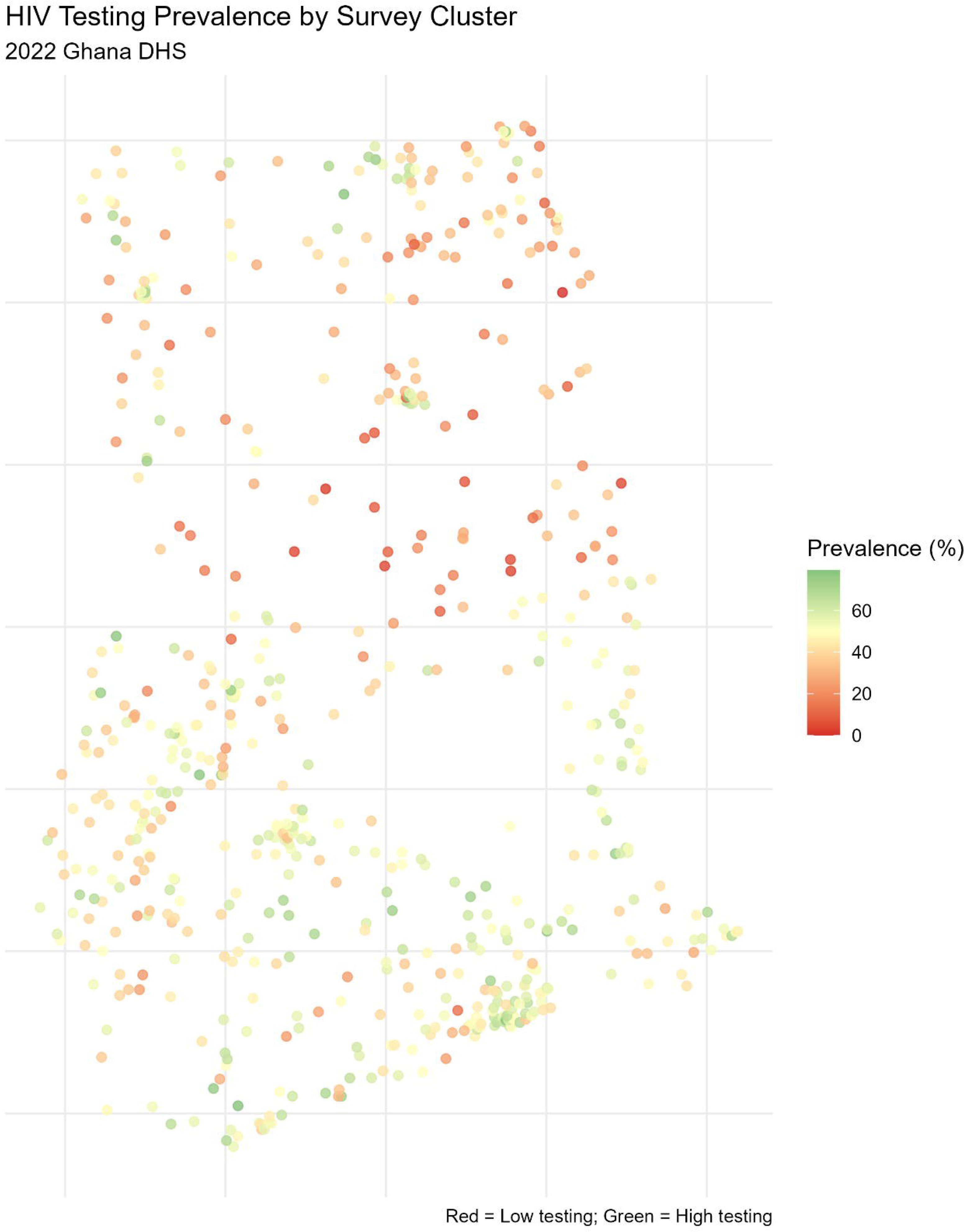
HIV Testing Prevalence by Survey Cluster, 2022 Ghana DHS. Each point represents one of the 618 DHS survey clusters. Red clusters indicate low testing prevalence; green clusters indicate high testing prevalence. Cluster GPS coordinates have been displaced by up to 2 km (urban) and 5 km (rural) to protect respondent confidentiality.

The 42 statistically significant High–High clusters (hotspots) and 65 Low–Low clusters (cold spots), along with a number of spatial outliers (six High–Low and 10 Low–High spatial outliers) were identified by the LISA analysis. No statistically significant local spatial clustering was observed among the 495 clusters that were not included in the first group. These clusters were geographically distributed and showed a clear spatial pattern. The hotspots were mainly located in southern Ghana, mainly in the Greater Accra, Eastern and Volta regions. Conversely, cold spots were mainly found in the Savannah, Northern and North East areas which aligned closely with the areas with the lowest HIV testing prevalence in the descriptive analysis. A north–south gap in uptake of HIV testing observed suggests that geographic inequalities in HIV testing are closely linked to other inequalities in socioeconomic development, health infrastructure, and service access in the country (Figure 4).

**Figure 4.**
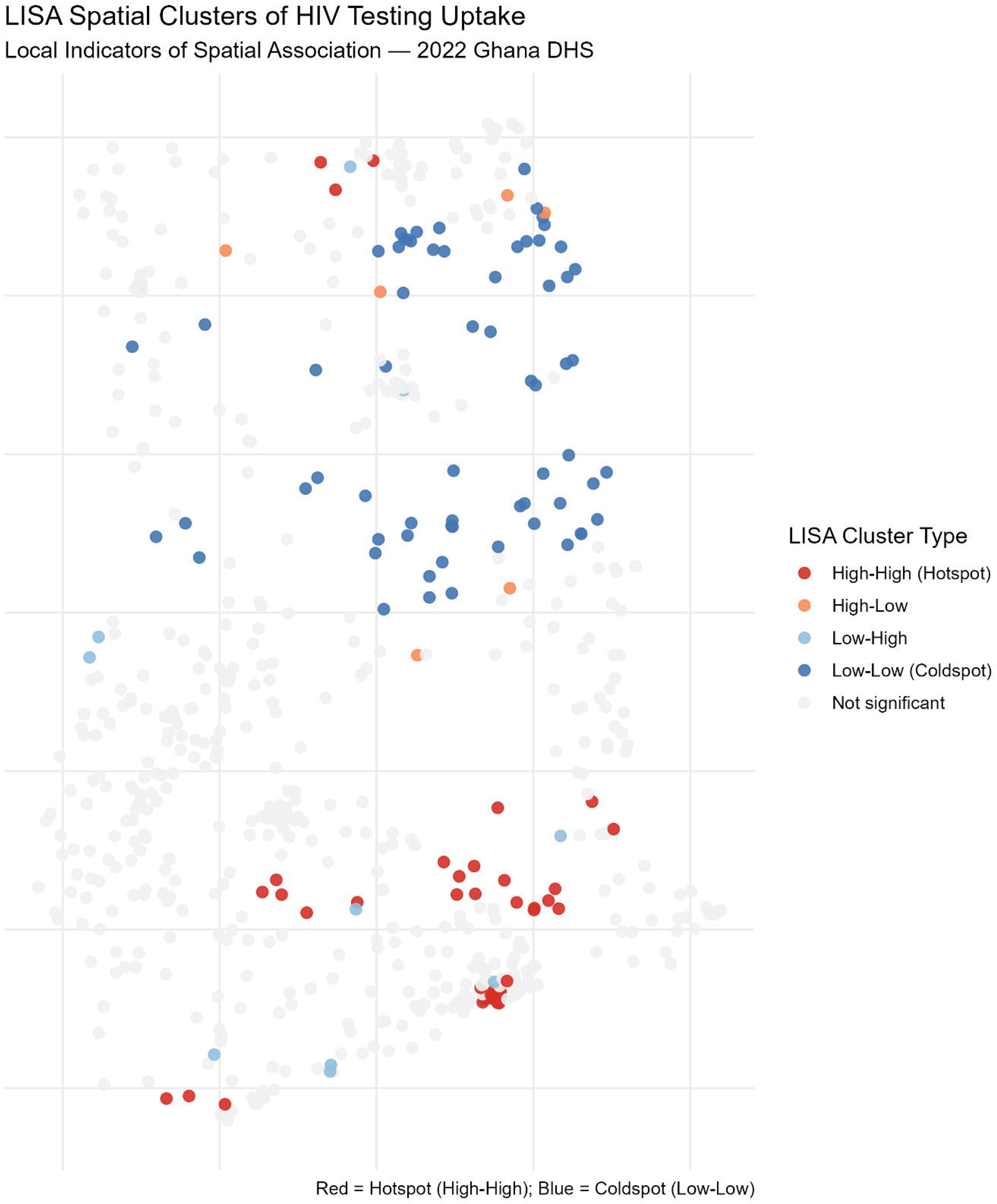
LISA Spatial Clusters of HIV Testing Uptake, 2022 Ghana DHS. High-High clusters (hotspots, red) are concentrated in southern Ghana; Low-Low clusters (coldspots, blue) are concentrated in northern Ghana. High-Low and Low-High clusters represent spatial outliers. Non-significant clusters are shown in grey.

### 3.4 Multilevel Logistic Regression

The intraclass correlation coefficient (ICC) for the null model was 0.052, suggesting that 5.2% of the variation in HIV testing uptake was due to differences between the survey clusters. While the clustering was small, it does indicate that there are some contextual features that shape HIV testing behaviour that are shared among people living in the same communities. There was a considerable inter-cluster variation that justified the use of a multilevel modelling approach. Adopting all the other covariates in the model, household wealth was strong and independent predictor of HIV uptake testing. HIV testing was found to be progressively higher among persons as their socioeconomic status increased, suggesting an increasing socioeconomic gradient for HIV testing. The odds of seeing HIV testing in the past were over two times higher in the richest quintile than in the poorest quintile. The overall trend of an increase in the odds regardless of wealth groups further demonstrates the results from the concentration index analysis and confirms that individuals with more socioeconomic resources use HIV testing services more than those with less socioeconomic resources. Education was a key factor associated with HIV testing. HIV testing rates also rose with each additional level of education, and individuals with higher education had significantly higher HIV testing rates compared to those with no education. But this correlation is strong, indicating that education could prove to be a factor in HIV testing in multiple ways: by increasing health literacy, awareness of HIV prevention services, decision-making ability, and access to health care. Pronounced sex differences were also evident. HIV testing was significantly lower among males compared to females, regardless of socioeconomic and demographic factors. This is consistent with the descriptive results and suggests that the gender disparity in HIV testing is not entirely attributable to disparities in wealth, education or locality. This increased uptake among women may be due to routine opportunities for HIV testing that are available during the provision of antenatal and other reproductive health services and are less frequently available for men. HIV testing uptake was strongly correlated with age. The risk of testing was much higher across all age groups than in adolescents aged 15-19 years and peaked in early and middle adulthood (30-34 years). The association was weaker in older age groups but was still consistently higher than in adolescents for all adult age groups. This trend indicates that HIV testing is more prevalent in the life stages of marriage, childbearing, and engagement in health care services. Another key predictor of HIV testing uptake was having a health insurance plan. People with health insurance were over twice as likely as people without health insurance to have been tested for HIV. This is an important clue to the potential importance of health care access and health care utilisation in influencing testing behavior. Those who are frequent users of health services might be more likely to receive HIV-related counselling, screening recommendations and referral services. HIV testing uptake was also linked to marital status. Never married people were significantly less likely to be tested for HIV than people currently married or in a relationship with a partner. This trend was also seen among former spouses, though the effect was less strong. These results could be attributed to different levels of perceived HIV risk, healthcare seeking behavior, and exposure to HIV testing opportunities experienced by marital relationships. Interestingly, the urban–rural difference found in the descriptive analysis was no longer significant after controlling for socioeconomic and demographic factors. Rural residence was not independently linked to HIV testing uptake, indicating that the lower prevalence in rural areas likely reflects the fact that there are differences in wealth, education, and insurance status. Similarly, after controlling for other socioeconomic factors, there was no significant association between regular exposure to radio or television and HIV testing, suggesting that the effect of media exposure might be mediated through other socioeconomic pathways. Overall, the multilevel analysis shows that socioeconomic position, education, gender, age, marital status and health insurance status are the most significant factors influencing HIV testing uptake in Ghana. The relationship between wealth and education inequality with access to and uptake of HIV testing services remained strong even after controlling for individual and community-level factors, highlighting the key role of structural inequalities (Table 4).

**Table 4.**
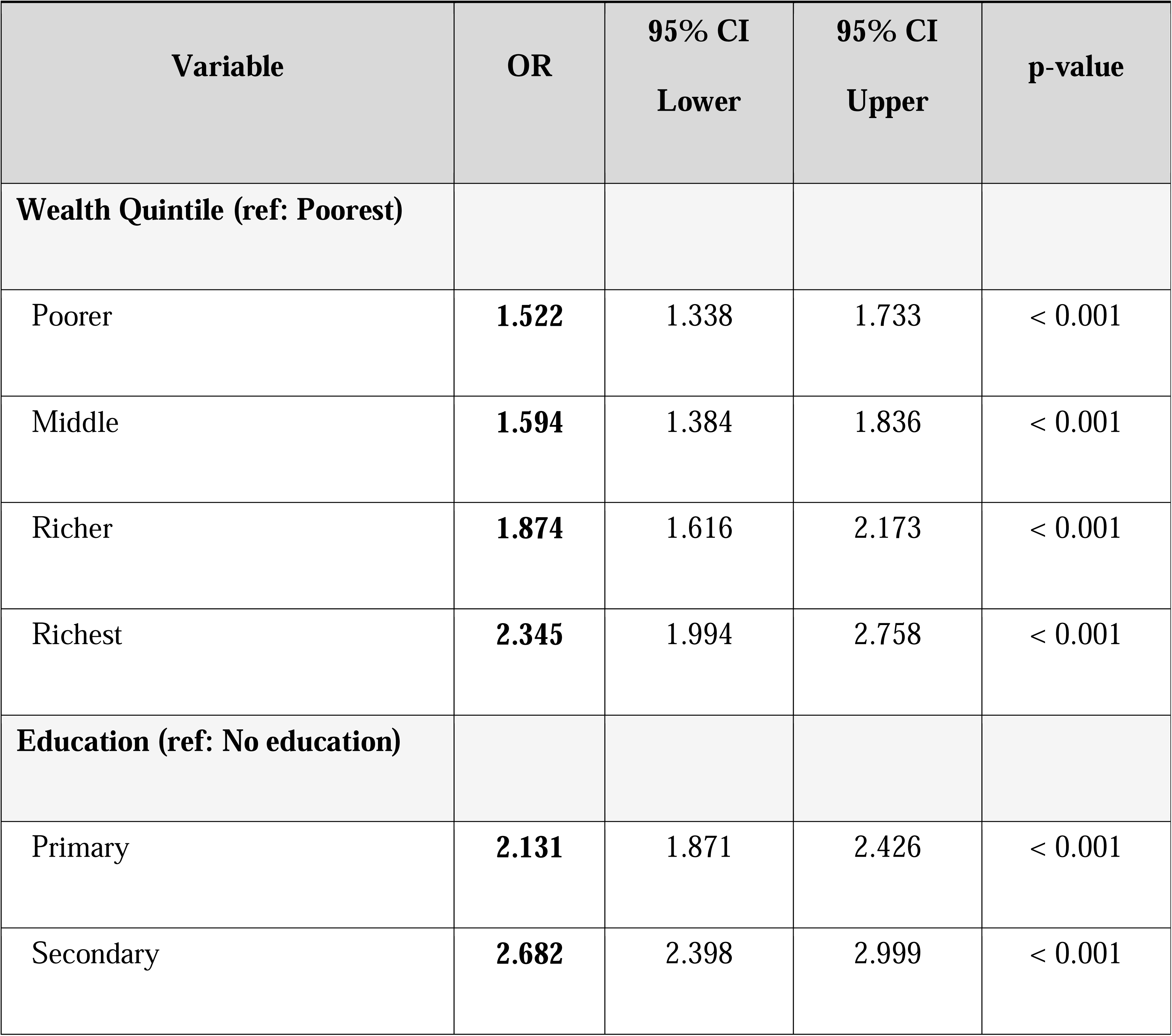
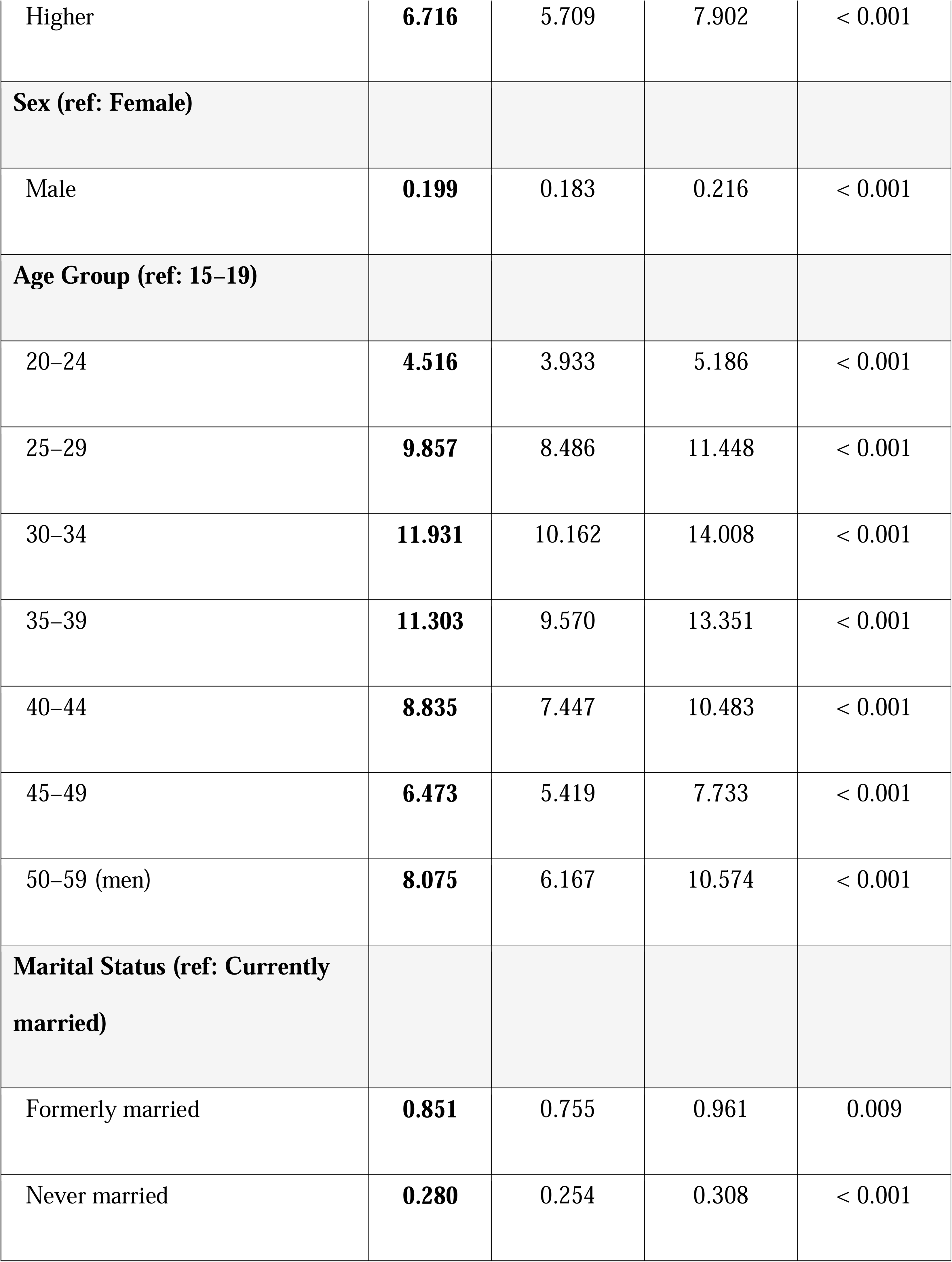
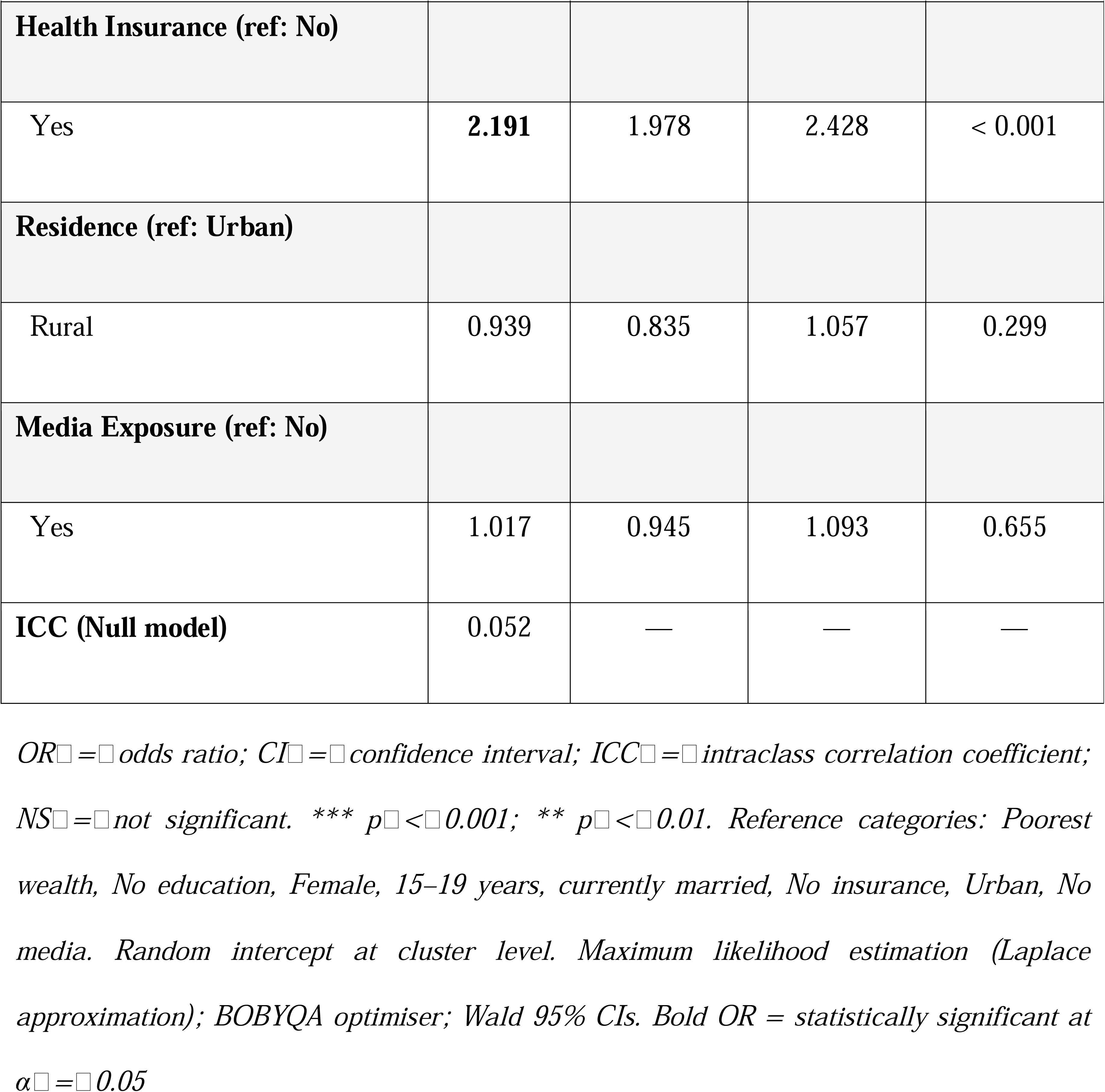
Multilevel Logistic Regression: Adjusted Odds Ratios for HIV Testing Uptake, 2022 Ghana DHS (n□=□21,754)

## 4. Conclusion

Therefore, the findings of this study indicated that HIV testing uptake in Ghana continued to have key socioeconomic and geographic disparities, despite free HIV testing services. The observed inequalities were concentrated among more highly educated and more affluent people, and wealth and education level were the main factors that accounted for these differences. Additionally, a spatial analysis showed that there were significant spatial clusters of HIV testing in southern Ghana and continued spatial cold spots in northern Ghana, highlighting the large geographic disparities in service use. The authors found that, after controlling for other factors, wealth, education, age, sex, marital status, and health insurance were independently associated with uptake of HIV testing, while rural residence and media exposure were not. The findings here suggest that the structural socioeconomic disadvantage remains to be a determinant of HIV testing equity. The disparities will have to be tackled with specific geographical interventions that will focus on disadvantaged groups, improve health system availability, and increase equitable access to HIV testing services in all parts of Ghana.

## Data Availability

This study was based on a cross-sectional secondary analysis of data obtained from the 2022 Ghana Demographic and Health Survey (GDHS).

https://dhsprogram.com

## Author Contributions

Abubakar Iddrisu Siddiq: conceptualization, methodology, data curation, formal analysis and writing of the original draft. Osman Abdul-Fatawu Iddrisu: data curation, formal analysis, writing, reviewing and editing. Abubakar Hudu Siddick: Methodology. Sebastina Naa Tawiah Sowah and Solomon Quansah Botchway: Writing, reviewing and editing of all sections. All authors read and approved the final manuscript.

